# Antigen rapid tests, nasopharyngeal PCR and saliva PCR to detect SARS-CoV-2: a prospective comparative clinical trial

**DOI:** 10.1101/2020.11.23.20237057

**Authors:** Jean Marc Schwob, Alix Miauton, Dusan Petrovic, Jean Perdrix, Nicolas Senn, Katia Jaton, Opota Onya, Alain Maillard, Gianni Minghelli, Jacques Cornuz, Gilbert Greub, Blaise Genton, Valérie D’Acremont

## Abstract

**Background:** Nasopharyngeal antigen Rapid Diagnostic Tests (RDTs) and saliva RT-PCR have shown variable performance to detect SARS-CoV-2.

**Methods:** In October 2020, we conducted a prospective trial involving patients presenting at testing centers with symptoms of COVID-19. We compared detection rates and performance of RDT, saliva PCR and nasopharyngeal (NP) PCR.

**Results:** Out of 949 patients enrolled, 928 patients had all three tests. Detection rates were 35.2% (95%CI 32.2-38.4%) by RDT, 39.8% (36.6-43.0%) by saliva PCR, 40.1% (36.9-43.3%) by NP PCR, and 41.5% (38.3-44.7%) by any test. For those with viral loads (VL) ≥10^6^ copies/ml, detection rates were 30.3% (27.3-33.3), 31.4% (28.4-34.5), 31.5% (28.5-34.6), and 31.6% (28.6-34.7%) respectively.

Sensitivity of RDT compared to NP PCR was 87.4% (83.6-90.6%) for all positive patients and 96.5% (93.6-98.3%) for those with VL≥10^6^. Sensitivity of STANDARD-Q^®^, Panbio™ and COVID-VIRO^®^ Ag tests were 92.9% (86.4-96.9%), 86.1% (78.6-91.7%) and 84.1% (76.9-89.7%), respectively. For those with VL≥10^6^, sensitivities were 96.6% (90.5-99.3%), 97.8% (92.1-99.7%) and 95.3% (89.4-98.5%) respectively. Specificity of RDT was 100% (99.3-100%) compared to any PCR. RDT sensitivity was similar <4 days (87.8%) and ≥4 days (85.7%) after symptoms onset (p=0.6). Sensitivities of saliva and NP PCR were 95.7% (93.1-97.5%) and 96.5% (94.1-98.1%), respectively, compared to the other PCR.

**Conclusions:** The high performance of RDTs allows rapid identification of COVID cases with immediate isolation of the vast majority of contagious individuals. RDT could be a game changer in primary care practices, and even more so in resource-constrained settings. PCR on saliva can replace NP PCR.

ClinicalTrial.gov Identifier: NCT04613310

## Introduction

COVID-19 is responsible for a dramatic health and social situation around the globe. Rapid and accurate detection of SARS-CoV-2 virus in symptomatic individuals is essential for taking immediate measures such as patient isolation and quarantine. Testing is the cornerstone of pandemic management.^1^ At present, nasopharyngeal (NP) swabbing followed by reverse transcription RT-qPCR analysis is the reference standard for detection of SARS-CoV-2 infection.^2^ This method requires trained staff to perform the swabbing, as well as laboratory personnel and sophisticated equipment to perform PCR analysis. In addition, NP swabbing causes discomfort for the patient and can lead rarely to complications.^3^ Also, the turnaround time for getting results is usually 24-48 hours, which forces most tested individuals to wait until they might resume usual activities if tested negative. Because of these limitations, there is definitely a need to investigate alternative testing methods to break transmission chains more rapidly, release the pressure on the health system and ease the way for patients.

To address the issues of laboratory infrastructure and procedures, as well as turnaround time, several companies have developed point of care antigen rapid diagnostic tests (RDT) to detect SARS-CoV-2. These tests are performed on NP swabs for the time being. Manufacturers report analytical sensitivities above 95% for all of these tests, while independent laboratory based studies report variable performances.^4,5^ However, very few studies have been performed in the so-called real world, i.e. in clinical settings where these tests will be applied. A recent study reports a clinical manufacturer-independent evaluation on three different RDT brands. Results show that sensitivity of the best-performing test (STANDARD Q^®^) was 77% and specificity 99%.^6^

To address the swabbing issue, there has been several attempts to use saliva for the detection of SARS-CoV-2, as it has been done for different viruses, including coronaviruses responsible for SARS and MERS.^7^ For SARS-CoV-2, a systematic review published on studies conducted up to April 2020 documented a sensitivity of 91% (95%CI 80–99%) for saliva and 98% (89–100%) for NP RT-PCR in previously confirmed COVID-19 patients.^8^ More recent studies, conducted in hospitalized patients, reported a sensitivity between 80%,^9,10^ and 100%.^11,12^

To simultaneously investigate analytical (PCR and RDTs) and sampling procedures (saliva and NP swab), we conducted a prospective clinical trial in symptomatic patients, in order to compare the detection rate of SARS-CoV-2 and sensitivities of i) RDT on NP swab, ii) PCR on NP swab and iii) PCR on saliva. Secondary objectives were to compare detection rates and sensitivities stratified by VL categories.

## Methods

### Ethic statement

The study protocol and related documents were approved by the ethical review committee of Canton Vaud (CER-VD 2020-02269).

### Brand of Rapid Diagnostic Tests evaluated

Three antigen-based RDTs were assessed: 1) STANDARD Q^®^ COVID-19 Ag Test from Biosensor/Roche, 2) Panbio™ COVID-19 Ag Test from Abbott and 3) COVID-VIRO^®^ from AAZ-LMB. All assays are lateral flow tests which detect viral nucleocapside antigens with color change assessed by naked eye reading. All tests were performed according to manufacturers’ information. RDT brands were rotated after around 30 positive patients until at least 100 positive per test were reached.

### SARS-CoV-2 RT-PCR, cycle thresholds and viral load quantification

SARS-CoV-2 RT-PCR were performed using an in-house RT-PCR on the automated molecular diagnostic platform targeting the E gene,^13–15^ or using the SARS-CoV-2 test of the Cobas 6800 instrument (Roche, Basel, Switzerland). Viral load were obtained by converting cycle thresholds of the RT-PCR instruments, using the formula logVL=-0.27Ct+13.04, as previously reported.^16,17^

### Outcomes

The primary outcome was the proportion of SARS-CoV-2 positive patients for PCR on saliva, PCR on NP swab, and RDT on NP swab. The secondary outcome were the viral loads of SARS-CoV-2 measured by PCR on saliva and NP swabs.

### Study design and participants

This was an observational prospective comparative clinical trial. Patients above 18 years were recruited from three different outpatient clinics in Lausanne with symptoms compatible with COVID-19 according to regional testing criteria (www.coronacheck.ch) (see supplementary file for details).

### Study procedures

Patients collected themselves the saliva for PCR analysis (swabbing their oral mucosa and finishing by drooling saliva in a tube) (see supplementary file for more details). Then, the health professional collected two nasopharyngeal swabs, one for PCR and one for RDT analyses. Test and control lines were read by the person having collected the swab after 15 to 20 minutes and judged as positive intense, positive weak, or negative. Remaining samples (saliva and second NP swab) were sent to the molecular diagnostics laboratory for RT-PCR analysis.

### Sample size

We based our sample size of 250 positive among 1250 cases tested to have a precision of ±2% on the detection rate if the latter was 20%.

### Statistical analysis

All patients having a result available for the 3 tests were included in the study analysis population. Detection rate, sensitivity, and specificity of each test with 95% confidence intervals (CIs) were estimated. Chi-square test was used for comparison between proportions, and Student t-test along with ANOVA F-tests for comparisons between log viral loads. Analyses were stratified by viral load categories. The thresholds chosen for binary stratified analyses by VL were 10^5^ copies/ml (Ct=30) and 10^6^ copies/ml (Ct=26), based on recent and older data investigating the link between viral loads and the presence of culture-competent virus.^18–22^

## Results

Presumed SARS-CoV-2 patients were screened from September 25^th^ to November 4^th^ 2020. 949 patients providing consent were enrolled. Median age was 31 years (IQR 25-42; range 18-87) with 49% being female. On the day of testing, 96% of participants had at least one major symptom (41% fever, 64% cough, 62% sore throat, 32% anosmia/ageusia) and 4% at least one minor and a close contact with a documented COVID-19 case. Mean duration of symptoms at the time of swab collection/testing was 2.6 days (SD 2.3, range 0-30).

Among the 928 who had all three tests done, 333 (36%) were tested using STANDARD Q^®^ COVID-19 Ag, 271 (29%) using the Panbio™ COVID-19 Ag, and 324 (35%) using the COVID-VIRO^®^ (see supplementary figure 1 for details on study flow).

### Detection rates of RDT, NP PCR and saliva PCR

Of the 928 patients analyzed, 327 (35.2%; 95%CI 32.2-38.4%) tested positive by RDT, 369 (39.8%; 36.6-43.0%) by saliva PCR, 372 (40.1%; 36.9-43.3%) by NP PCR, and 385 (41.5%; 38.3-44.7%) by any of the 3 tests (Figure 1A). Detection rates were thus equivalent for both NP and saliva PCR (p=0.9), with NP PCR detecting 16 additional cases when compared to saliva PCR, and saliva PCR detecting 13 additional cases when compared to NP PCR. The detection rate for RDT was significantly inferior to NP PCR (p=0.03) and saliva PCR (p=0.04), with NP and saliva PCR detecting 45 and 42 additional cases compared to RDT, respectively.

**Figure 1:**
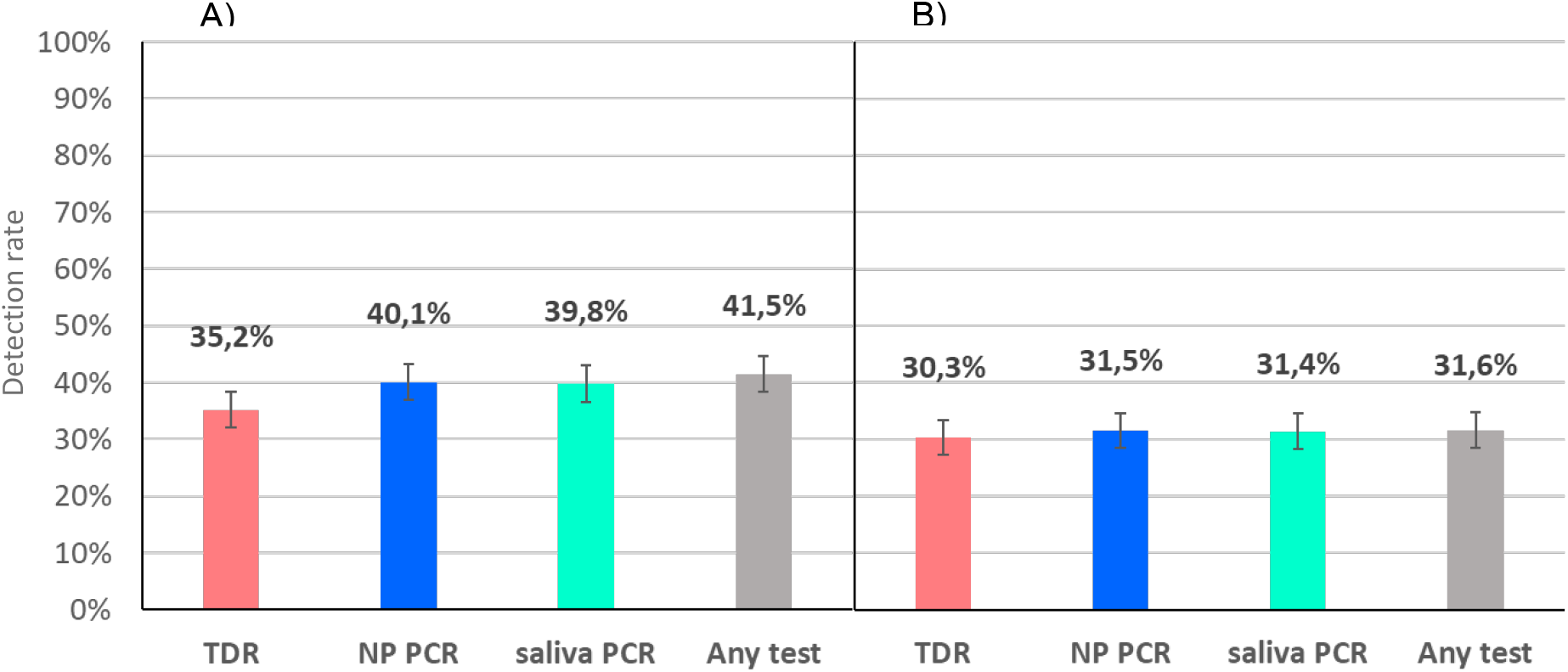
Detection rates of COVID patients by RDT, NP PCR and saliva PCR: A) all positive patients; B) positive patients with viral loads ≥10^6^ copies/ml by any PCR (supposedly significantly contagious)

When considering the 293 (31.6%) patients with a VL ≥10^6^ copies/ml by either NP or saliva PCR, the differences were lower with a detection rate of 30.3% (27.3-33.3) (281/928) for RDT, 31.4% (28.4-34.5) (291/928) for saliva PCR, 31.5% (28.5-34.6) (292/928) for NP PCR, and 31.6% (28.6-34.7%) by any test (Figure 1B). There were no more significant differences between detection rates of PCRs and RDT (p=0.6).

### Diagnostic test performance (sensitivity, specificity) of RDTs, NP PCR and saliva PCR

The sensitivity of RDT compared to NP PCR was 87.4% (83.6-90.6). When considering those with a VL ≥10^6^ copies/ml by NP PCR, sensitivity was 96.5% (93.6-98.3%) (Figure 2). The sensitivity of RDT according to viral load categories remained higher than 95% up to 10^6^ and dropped rapidly for lower loads (Figure 3). Sensitivities of the three RDT brands were 92.9% (86.4-96.9%) for STANDARD Q^®^, 86.1% (78.6-91.7%) for Panbio™ and 84.1% (76.9-89.7%) for COVID-VIRO^®^ Ag tests (p=0.1). When considering those with a VL ≥10^6^ copies/ml by NP PCR, sensitivities were 96.6% (90.5-99.3%), 97.8% (92.1-99.7%) and 95.3% (89.4-98.5%) respectively (Figure 2). Of note, the median log VL of all participants were similar between the 3 brands of RDT (Supplementary Figure 3).

**Figure 2:**
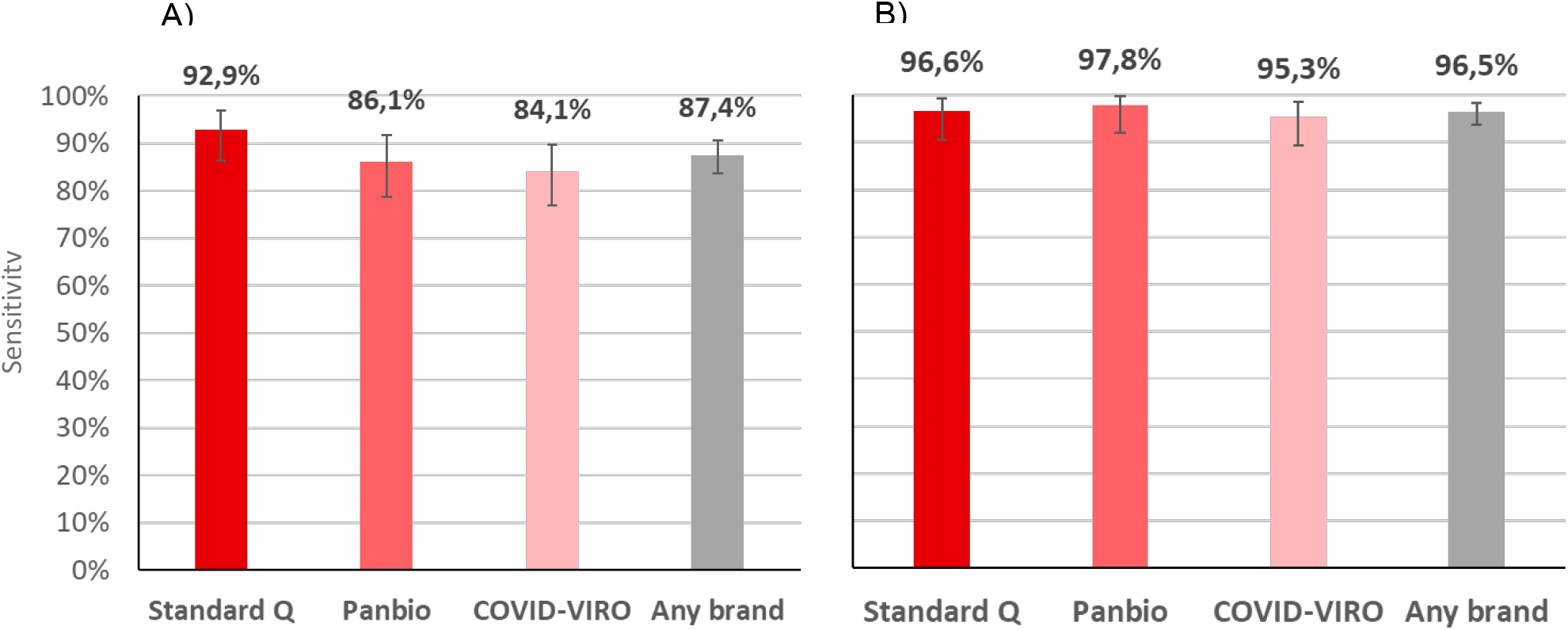
Sensitivity of three brands of antigen RDT compared to NP PCR: A) all positive patients; B) positive patients with viral loads ≥10^6^ copies/ml (supposedly significantly contagious)

**Figure 3:**
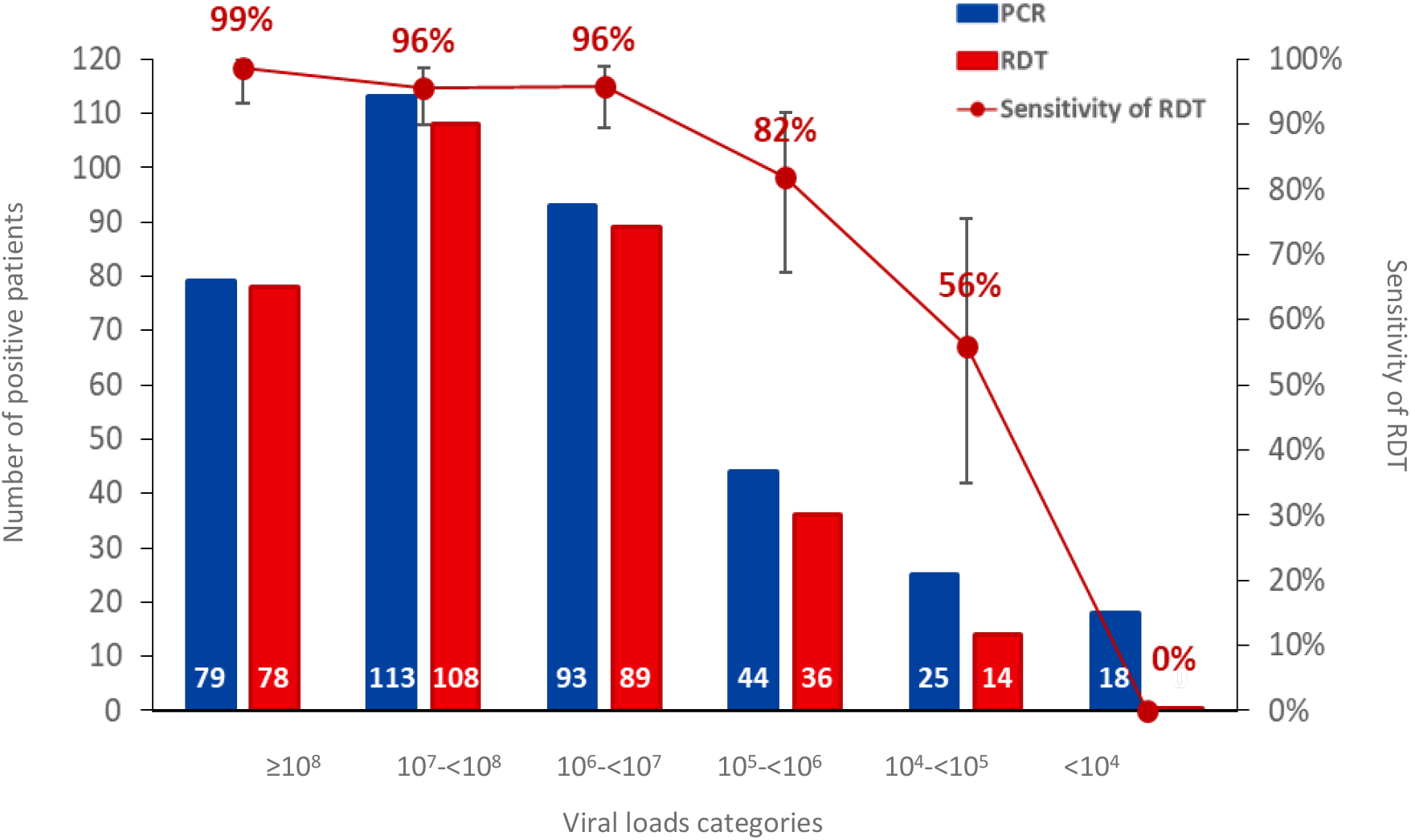
Number of patients positive by RDT (in red) and NP PCR (in blue) and sensitivity of RDT according to viral load categories

The diagnostic performance of NP PCR and saliva PCR were equivalent: sensitivities were 95.7% (93.1-97.5%) for saliva compared to NP PCR and 96.5% (94.1-98.1%) for NP compared to saliva PCR.

When using the composite reference standard (385 patients with any test positive), sensitivities were 84.9% (81.0-88.3%) for RDT, 95.8% (93.3-97.6%) for saliva PCR, and 96.6% (94.3-98.2%) for NP PCR. The difference was not significant between NP PCR and saliva PCR (p=0.6), but was so between RDT and NP PCR or saliva PCR (p<0.001). Specificity of RDT was 100% (99.3-100%), which means that there was no false positive RDT. In two instances, RDT was positive and NP PCR was negative, but the saliva PCR was positive with viral loads of 4.0×10^8^ and 1.7×10^3^.

### Virus loads (VL)

VLs of patients with positive saliva PCR (median 1.3×10^5^; IQR 1.9×10^4^-8.8×10^5^; range 2.0-9.3) were significantly lower than those with positive NP PCR (median 1.3×10^7^; IQR 1.3×10^6^-8.4×10^7^; range 2.0-9.4) (p<0.001). Among 285 patients with VL≥10^6^ by NP PCR, 10 (3.5%) were negative by RDT. Among 87 patients with VL<10^6^ by NP PCR, 50 (57.5%) were positive by RDT (Figure 4A). The difference between NP and saliva VLs is illustrated in Figure 4B.

**Figure 4:**
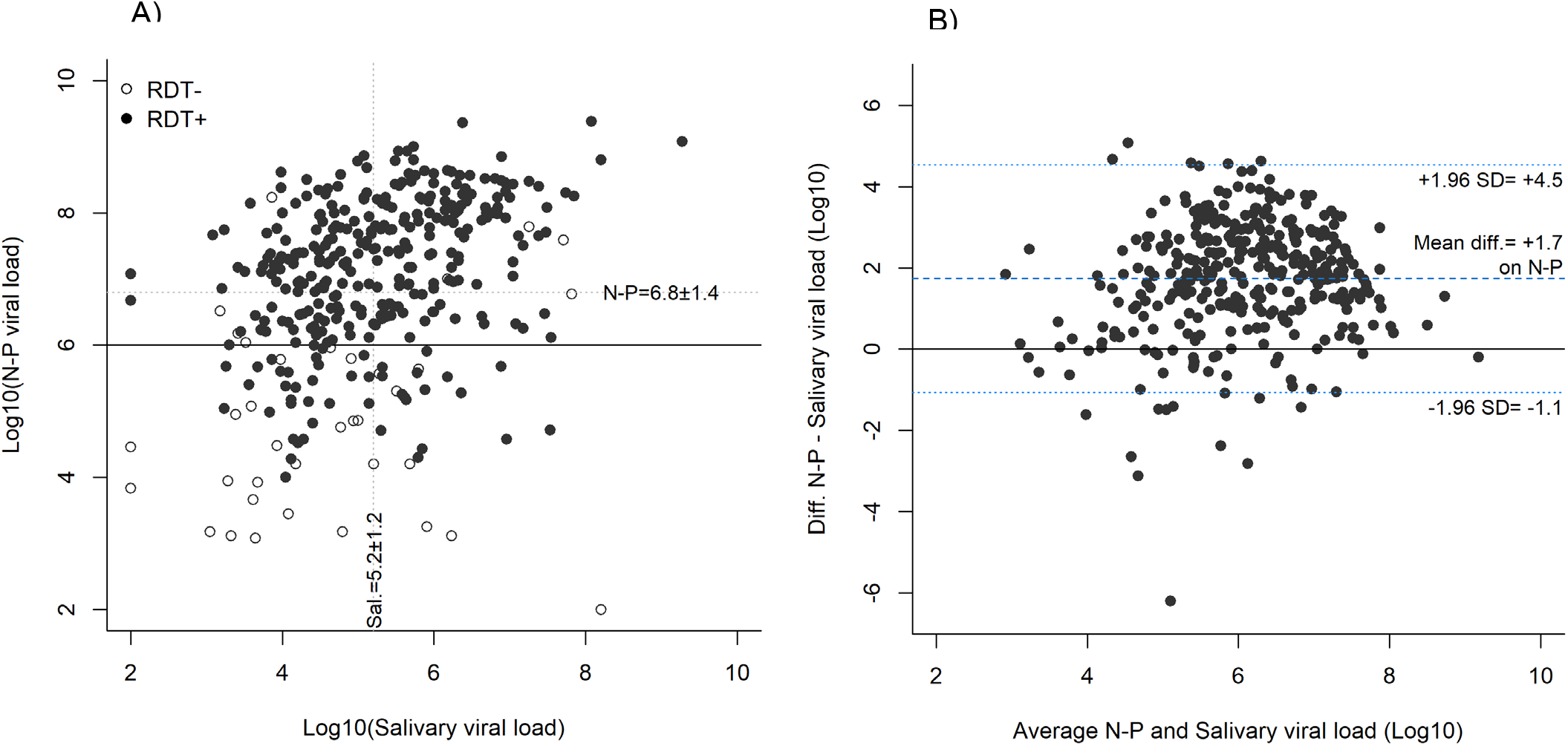
Comparison between log viral loads by NP PCR and salivary PCR; A) Log viral loads in RDT positive (black dots) and negative (white dots) patients. Dotted lines: mean log viral loads; Black line: considered threshold for presence of cultivable virus (NP PCR); B) Bland-Altman analysis showing the difference between NP and saliva log viral loads; SD=Standard deviation.

The sensitivity of RDT according to symptoms duration varied between 80% and 90%. It was lowest the day of symptoms onset (80%, 95%CI 44.4-97.5%) and highest on day 4 (90.0%, 73.5-97.9%) (Figure 5). There was no significant difference in the sensitivity of RDT, neither between the first 4 days of symptoms (87.8%) and thereafter (85.7%) (p=0.6), nor the first 7 days (87.7%) and thereafter (81.3%) (p=0.5).

**Figure 5:**
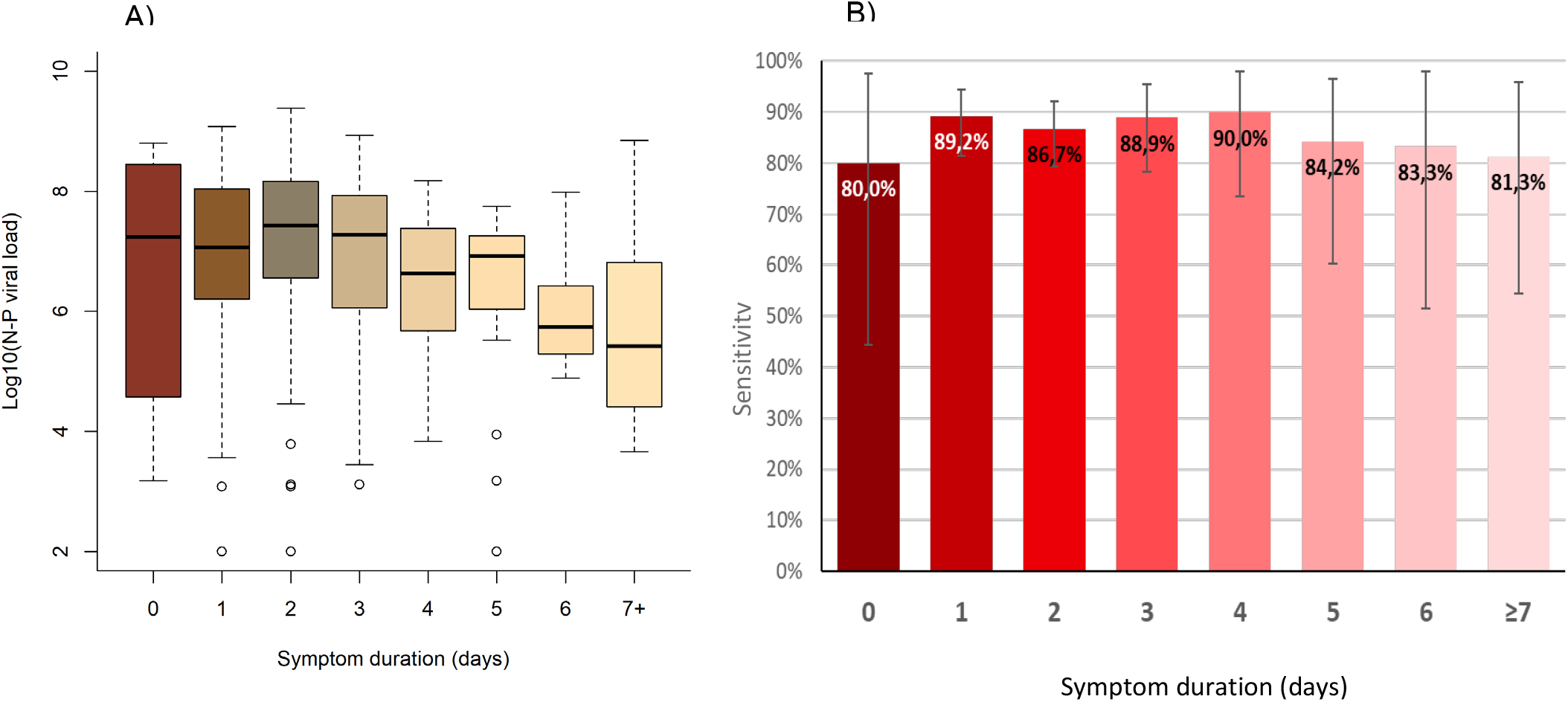
Log viral loads by NP PCR (A) and sensitivity of antigen RDT (B) according to symptoms duration

Comparison of VLs in RDT positive vs negative patients, as well as according to RDT color band intensity, type of symptoms and saliva volume are detailed in the supplementary file and supplementary figures.

## Discussion

The results of the present study show that the detection rate of positive COVID-19 cases by RDT was high, especially for those with a VL of ≥10^6^ copies/ml. The sensitivity of RDT compared to NP PCR was above 85%, and above 95% for patients with a high VL, and was similar between the three RDT brands, with an advantage for Standard Q^®^. The sensitivity varied slightly according to symptoms duration but remained above 80%, even after 7 days despite a progressive drop in VLs.^6^ Actually, a lower sensitivity after the acute phase of disease might be an advantage to prevent unnecessary isolation of patients who are, for most of them, no more contagious, despite a positive PCR result (known to sometimes persist for a long time).^23,24^ Our results are quite strong since they are based on a high number of positive patients and two PCR performed in parallel on each patient. We reached 385 positive cases, with >100 by RDT brand, thus fulfilling the FIND/WHO requirements for validating rapid diagnostic tests.

The detection rate of SARS-CoV-2 by PCR performed on a saliva sample was equivalent to that of RT-PCR performed on NP swabs. The sensitivity of PCR of one type of sampling compared to the other were similar and above 95%. The two positive saliva PCR but negative NP PCR patients who were still detected by RDT illustrates that some but rare false negative results of tests based on NP swabs are likely due to sampling procedure.

### RDT versus PCR

The sensitivity of RDT of more than 95% in patients with VL≥10^6^ copies/ml means that these rapid tests are likely to identify reliably individuals that are contagious, which would, if largely deployed, reduce transmission more substantially than what would be expected from its imperfect overall sensitivity. Furthermore, the short turn-around time to return the result to the patient allows more rapid isolation of cases and efficient contact tracing, which should also contribute to more efficient pandemic control.

There was a slight variability in performance between the three different RDTs with STANDARD Q® having a higher sensitivity (93%) than those of Panbio™ (86%) and COVID-VIRO® (84%), but all met the threshold of 80% sensitivity and 97% specificity of the WHO recommendations for use^25^. The sensitivities of all tests in the present study were higher than those reported in the first manufacturer-independent clinical validation study.^6^ The reasons might be that Krüger *et al* included a more diverse subject population, with a small number of positive samples (70), and used different PCR targets.^6^ Our high number of patients investigated with well-defined conditions and standardized procedures make results of the present work rather robust.

The specificity of all three tests was 100%, which is impressive considering the potential for inter-observer variation in RDT test line reading. This observation implies that the assessed RDTs brands are easy to read, and that faint lines can still be easily detected. This perfect specificity, which was also shown in the other studies on high quality RDT, allows to state that there is no need to confirm a positive RDT test result by an additional PCR test.

One of the strength of our study also lies in the fact that the study population represented that of routine COVID-19 diagnostic centers, namely symptomatic outpatients with fever or cough or anosmia/ageusia or symptomatic close contacts. The study was performed at the end-user level in real-life conditions, which is in agreement with WHO recommendations for evaluating the performance of new diagnostic tests.^26^ Real-life evaluation also allows to take into account context-specific factors that could influence clinical accuracy, such as common comorbidities.

### Saliva PCR versus NP PCR

Having a detection rate of saliva PCR equivalent to that of NP PCR is in line with a previous study done on 70 patients COVID-19 positive that showed excellent concordance between the two sampling methods.^12^ Other studies showed much lower performance with around 80% sensitivity for saliva PCR when compared to NP PCR. These divergent results are likely to be due to the sampling procedure and inhibitors that can interfere with amplification.

The median SARS-CoV-2 VL in saliva was approximately two log lower than that in the NP swab. With such a difference, an overall lower sensitivity of saliva PCR when compared to NP PCR would have been expected. This was not the case in the present evaluation because it is essentially the peak and not the extremes of the VL distribution curve that is shifted towards a lower value for saliva.

In terms of procedures, patients were able to easily perform the saliva sampling on themselves after getting a precise explanation by the health professional. Some were not able though to drool saliva in the tube, but this did not affect the sensitivity, the VL being not significantly lower in this group (supplementary figure 4).

The FDA has granted emergency use authorization to two saliva-based assays for the SARS-CoV-2, namely Rutgers’ RUCDR Infinite Biologics, and Curative-Korva SARS-Cov-2 Assay. Both of these assays are now in use to test for COVID-19, in spite of the fact that no independent scientific analysis has yet established their effectiveness. Our pragmatic approach, using the same transport medium tube as for NP swabs, can be applied in any testing facility. The similar sensitivity and specificity achieved by sampling the saliva instead of the nasopharynx validates the sampling method and procedure, at least in this outpatient population with relatively high viral load. The results of the present study, together with that of Wyllie *et al*,^12^ provide evidence that RT-PCR on saliva can be used for SARS-CoV-2 detection to ease testing and improve comfort and safety.

### Clinical significance

If RDT would be used in settings with a lower SARS-CoV-2 prevalence, such as 10%, a negative test would have a negative predictive value (NPV) of 98.6%. Such an NPV is acceptable if the patients do not belong to high-risk populations (severe cases, hospitalized patients). If the prevalence would be only 1%, the NPV would be 99.9%. Other NPV, whenever for RDT or PCR, can be simply calculated using the following formula: (1-P)/1-SP (P=prevalence; S=sensitivity), the specificity being 100%.

Regarding the positive predictive value (PPV), even taking the lowest specificity confidence interval (99.3%), it would be 93% at 10% prevalence, which is high enough. At a lower prevalence, the PPV would drop and the solution would be to restrict testing to patients with a high enough pre-test probability rather than confirming each positive case by PCR.

### Limitations

The present study was conducted in a well-defined outpatient population with usual testing criteria for COVID-19 and presenting within 7 days after symptom onset for most of them. Our results might not apply in a setting where patients would attend after one week of symptoms, keeping in mind that these outpatients would be much less likely to transmit.^27^ Our results might neither apply to hospitalized patients, who tend to present late in the course of the disease, thus with lower viral loads. We did not include children in the study; however, viral loads seem not to differ between children and adults,^28^ which suggests that RDT could perform similarly in younger age groups. We cannot infer the accurate diagnostic performance of saliva PCR and RDTs in an asymptomatic population that was not investigated here. The sufficient sensitivity (82%) in patients with viral loads between 10^5^ and 10^6^ suggests that RDTs can be safely used for screening schools, university students or contacts of SARS-CoV-2 positive patients. ^29^

## Conclusion

The very good performance of the best available RDTs allows point of care management with immediate relevant isolation of the majority of contagious individuals. The almost perfect concordance between saliva PCR and NP PCR results, the ease of administration and the safety of the procedure could trigger change in sampling method using saliva as reference standard, at least in outpatients who have higher viral loads than inpatients. RDT complies with the ASSURED (Affordable, Sensitive, Specific, User-friendly, Rapid and robust, Equipment-free and Deliverable to end-users) criteria, which makes them very useful in primary care practices, and even more so in resource-constrained settings.^30^ The variability in performance of different RDTs highlights the need to continue the efforts for having manufacturer-independent validation in the population that will be the target group for use, before implementation of a new brand at large-scale.

## Data Availability

All data referred to in the manuscript are available.

## Acknowledgements

We thank Yann Sancosme, Maxime Hostettler, Marion de Vallière and Maria Daniela Garrido for help in patient recruitment; Pierrette Meige, Catherine Mialet, Chantal Ngarambe, Tina Wyllie, Mani Souvannaraj, Annie Herard, Vincent Gliven, Maxime Naoux, Vania Carreira Augusto, Mélanie Crelier, Marie Jampen, Alain Lagacé and Jean-Luc Billaud of the testing centers for their work, the patients for consenting to have all samples collected and the health authorities of the canton de Vaud for their support. We thank René Brouillet, Marie-Anne Page, Zahera Naseri and all the team of the Laboratory of Molecular Diagnostics of the Institute of Microbiology.

---

**Supplementary figure 1:**
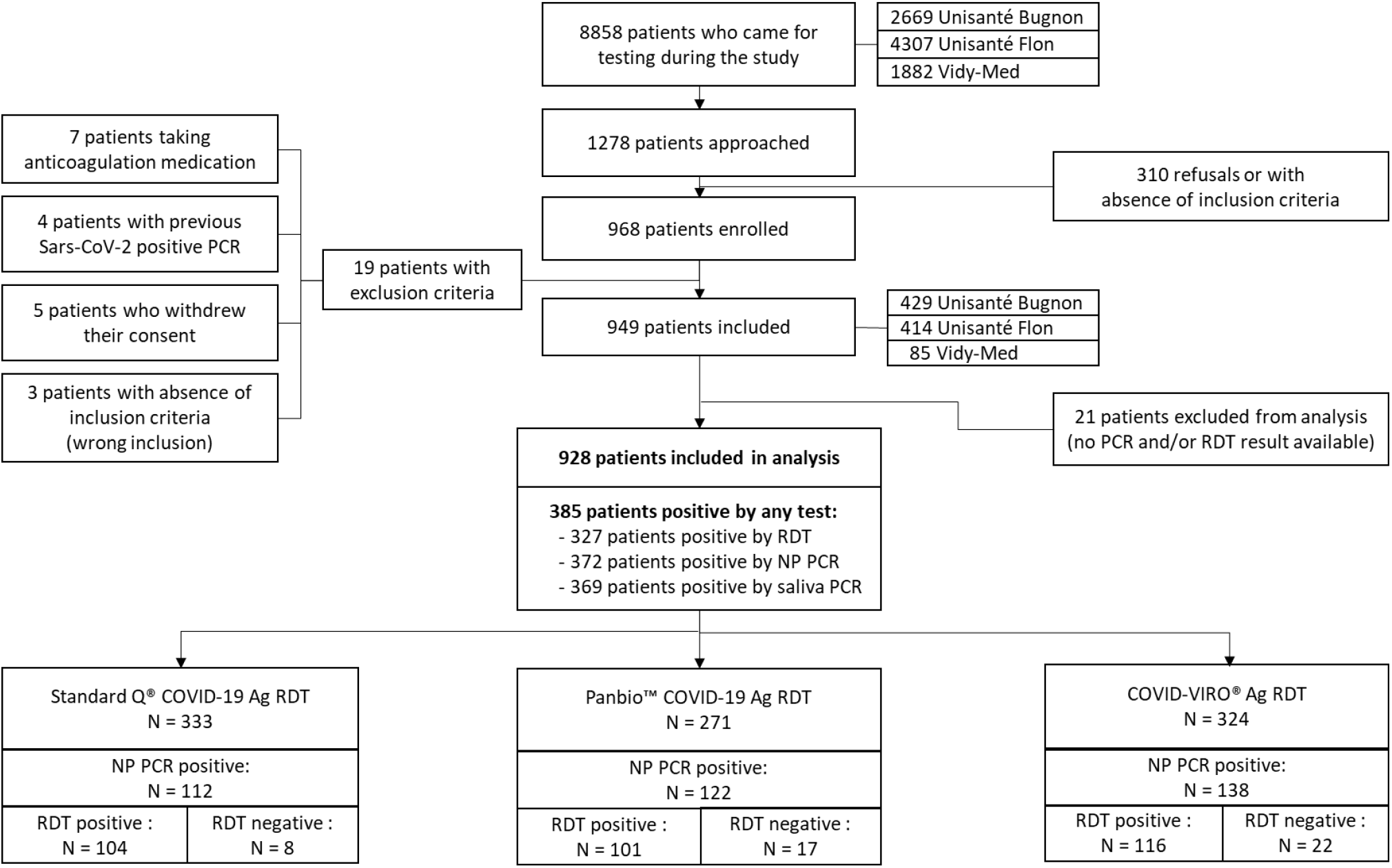
Study flow

**Supplementary figure 2:**
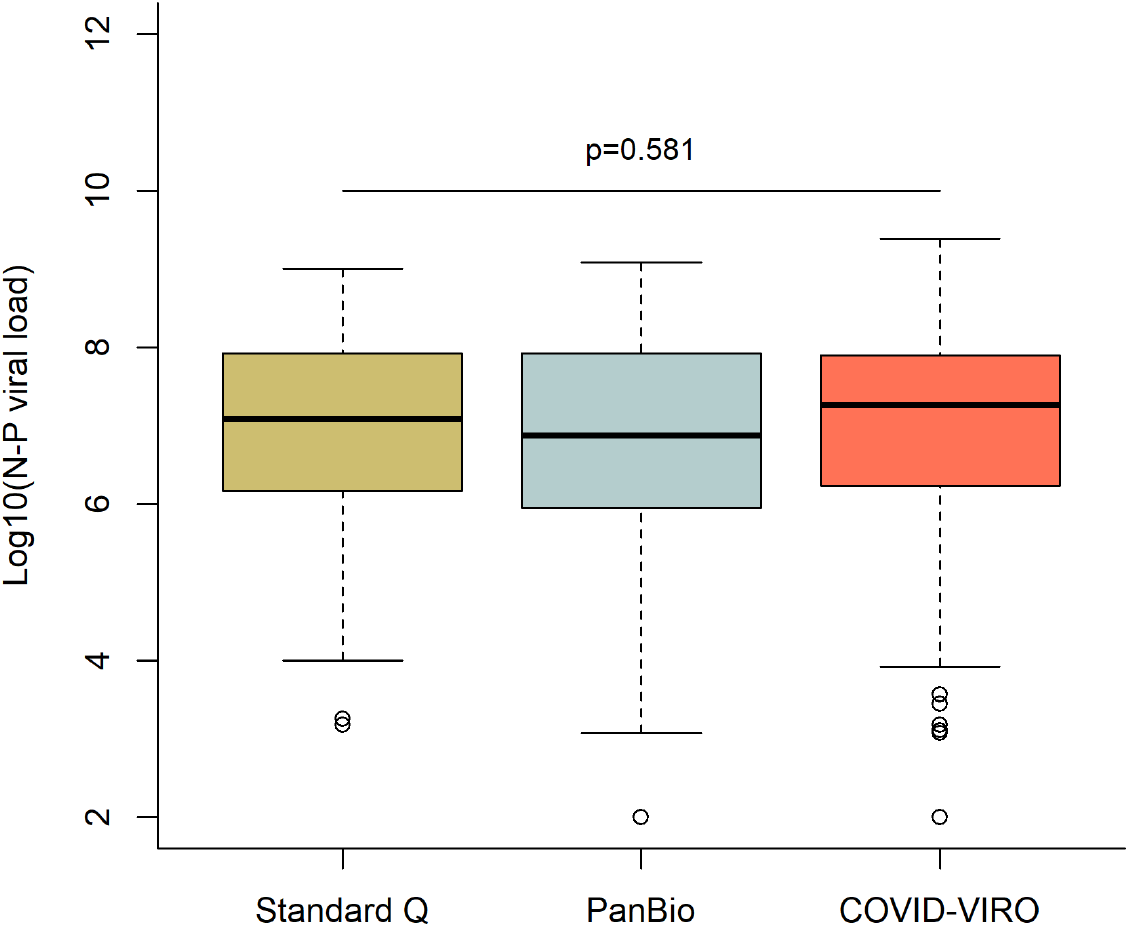
Log viral loads by NP PCR according to the RDT brand used.

**Supplementary figure 3:**
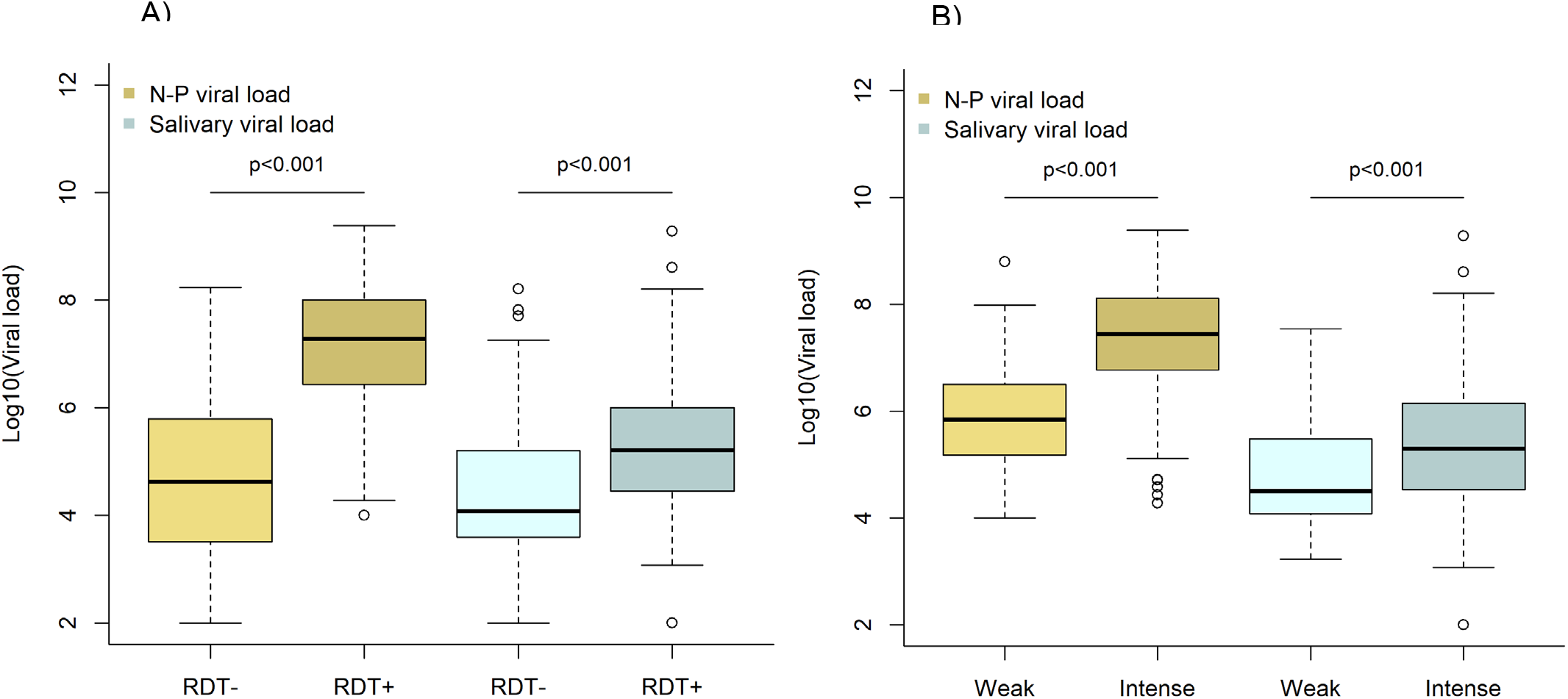
Log viral loads by NP PCR and saliva PCR according to A) RDT result and B) intensity band of positive RDT

**Supplementary figure 4:**
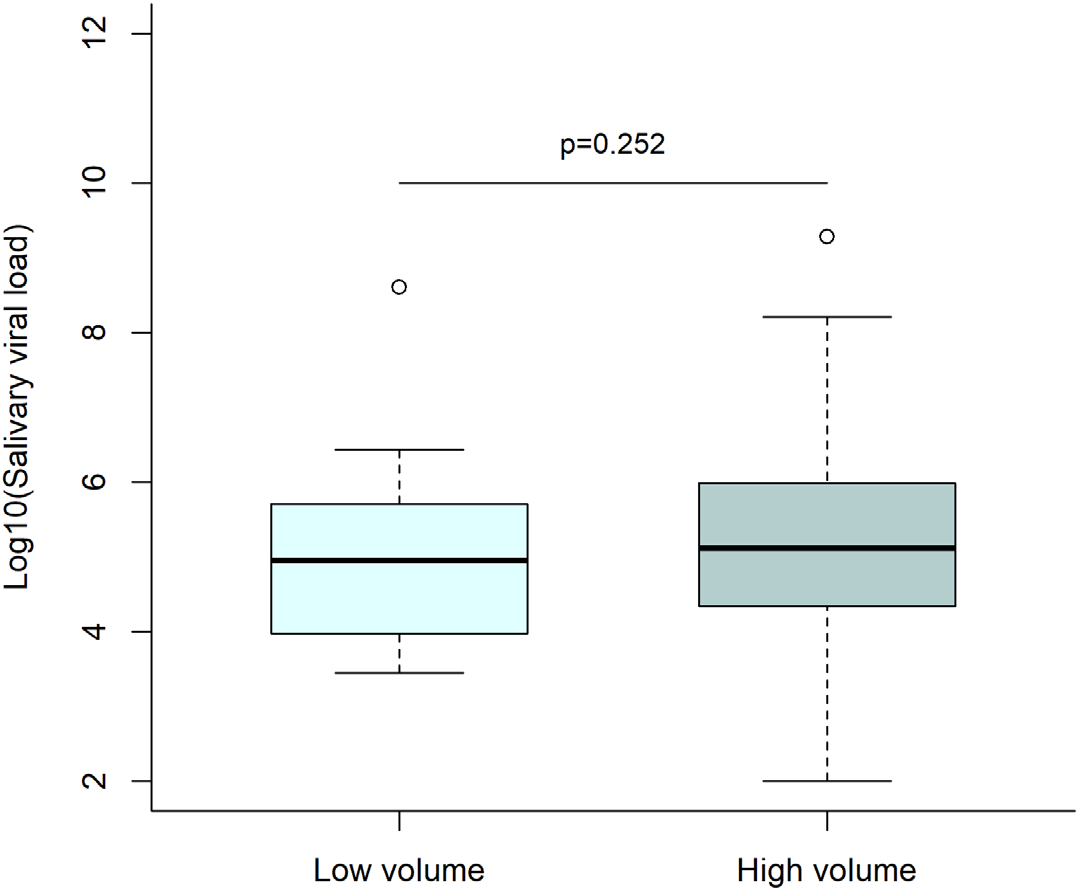
Log viral loads by saliva PCR saliva volume: low volume corresponds to a gingivo-buccal swab only; high volume corresponds to a gingivo-buccal swab with <0.5 ml of saliva in addition.

## References

1. Salath M, Althaus CL, Neher R, et al. COVID-19 epidemic in Switzerland: on the importance of testing, contact tracing and isolation. Swiss Med Wkly 2020;

2. Lippi G, Simundic A-M, Plebani M. Potential preanalytical and analytical vulnerabilities in the laboratory diagnosis of coronavirus disease 2019 (COVID-19). Clinical Chemistry and Laboratory Medicine 2020;58(7):1070–6.

3. Li L, Shim T, Zapanta PE. Optimization of COVID-19 testing accuracy with nasal anatomy education. American Journal of Otolaryngology 2021;42(1):102777.

4. Corman VM, Haage VC, Bleicker T, et al. Comparison of seven commercial SARS-CoV-2 rapid Point-of-Care Antigen tests [Internet]. 2020. Available from: <https://www.medrxiv.org/content/10.1101/2020.11.12.20230292v1preprint >

5. Dinnes J, Deeks JJ, Adriano A, et al. Rapid, point-of-care antigen and molecular-based tests for diagnosis of SARS-CoV-2 infection. Cochrane Database of Systematic Reviews 2020;

6. Krüger LJ, Gaeddert M, Köppel L, et al. Evaluation of the accuracy, ease of use and limit of detection of novel, rapid, antigen-detecting point-of-care diagnostics for SARS-CoV-2 [Internet]. 2020. Available from: <https://www.medrxiv.org/content/10.1101/2020.10.01.20203836v1preprint >

7. Niedrig M, Patel P, El Wahed AA, Schädler R, Yactayo S. Find the right sample: A study on the versatility of saliva and urine samples for the diagnosis of emerging viruses. BMC Infect Dis 2018;18(1):707.

8. Czumbel LM, Kiss S, Farkas N, et al. Saliva as a Candidate for COVID-19 Diagnostic Testing: A Meta-Analysis. Front Med 2020;7:465.

9. Altawalah H, AlHuraish F, Alkandari WA, Ezzikouri S. Saliva specimens for detection of severe acute respiratory syndrome coronavirus 2 in Kuwait: A cross-sectional study. Journal of Clinical Virology 2020;132:104652.

10. Nagura-Ikeda M, Imai K, Tabata S, et al. Clinical Evaluation of Self-Collected Saliva by Quantitative Reverse Transcription-PCR (RT-qPCR), Direct RT-qPCR, Reverse Transcription– Loop-Mediated Isothermal Amplification, and a Rapid Antigen Test To Diagnose COVID-19. J Clin Microbiol 2020;58(9):e01438–20.

11. Azzi L, Carcano G, Gianfagna F, et al. Saliva is a reliable tool to detect SARS-CoV-2. Journal of Infection 2020;81(1):e45–50.

12. Wyllie AL, Fournier J, Casanovas-Massana A, et al. Saliva or Nasopharyngeal Swab Specimens for Detection of SARS-CoV-2. N Engl J Med 2020;383(13):1283–6.

13. Corman VM, Landt O, Kaiser M, et al. Detection of 2019 novel coronavirus (2019-nCoV) by real-time RT-PCR. Eurosurveillance 2020;25(3).

14. Greub G, Sahli R, Brouillet R, Jaton K. Ten years of R&D and full automation in molecular diagnosis. Future Microbiology 2016;11(3):403–25.

15. Pillonel T, Scherz V, Jaton K, Greub G, Bertelli C. Letter to the editor: SARS-CoV-2 detection by real-time RT-PCR. Eurosurveillance 2020;25(21).

16. Jacot D, Greub G, Jaton K, Opota O. Viral load of SARS-CoV-2 across patients and compared to other respiratory viruses. Microbes and Infection 2020;in press.

17. Opota O, Brouillet R, Greub G, Jaton K. Comparison of SARS-CoV-2 RT-PCR on a high-throughput molecular diagnostic platform and the cobas SARS-CoV-2 test for the diagnostic of COVID-19 on various clinical samples. Pathogens and Disease 2020;78(8):ftaa061.

18. Bullard J, Dust K, Funk D, et al. Predicting Infectious Severe Acute Respiratory Syndrome Coronavirus 2 From Diagnostic Samples. Clinical Infectious Diseases 2020;ciaa638.

19. Jaafar R, Aherfi S, Wurtz N, et al. Correlation Between 3790 Quantitative Polymerase Chain Reaction–Positives Samples and Positive Cell Cultures, Including 1941 Severe Acute Respiratory Syndrome Coronavirus 2 Isolates. Clinical Infectious Diseases 2020;ciaa1491.

20. L’Huillier AG, Torriani G, Pigny F, Kaiser L, Eckerle I. Culture-Competent SARS-CoV-2 in Nasopharynx of Symptomatic Neonates, Children, and Adolescents. Emerg Infect Dis 2020;26(10):2494–7.

21. Singanayagam A, Patel M, Charlett A, et al. Duration of infectiousness and correlation with RT-PCR cycle threshold values in cases of COVID-19, England, January to May 2020. Euro Surveill 2020;25(32).

22. van Beek J, Igloi Z, Boelsums T, et al. From more testing to smart testing: data-guided SARS-CoV-2 testing choices [Internet]. 2020. Available from: <https://www.medrxiv.org/content/10.1101/2020.10.13.20211524v2preprint >

23. Bi Q, Wu Y, Mei S, et al. Epidemiology and transmission of COVID-19 in 391 cases and 1286 of their close contacts in Shenzhen, China: a retrospective cohort study. The Lancet Infectious Diseases 2020;20(8):911–9.

24. Lee S, Kim T, Lee E, et al. Clinical Course and Molecular Viral Shedding Among Asymptomatic and Symptomatic Patients With SARS-CoV-2 Infection in a Community Treatment Center in the Republic of Korea. JAMA Intern Med 2020;180(11):1447.

25. World Health Organization. Antigen-detection in the diagnosis of SARS-CoV-2 infection using rapid immunoassays: Interim guidance [Internet]. 2020. Available from: <https://www.who.int/publications/i/item/antigen-detection-in-the-diagnosis-of-sars-cov-2infection-using-rapid-immunoassays

26. Kosack CS, Page A-L, Klatser PR. A guide to aid the selection of diagnostic tests. Bull World Health Organ 2017;95(9):639–45.

27. Ferretti L, Ledda A, Wymant C, et al. The timing of COVID-19 transmission [Internet]. 2020. Available from: <https://www.medrxiv.org/content/10.1101/2020.09.04.20188516v2preprint >

28. Baggio S, L’Huillier AG, Yerly S, et al. SARS-CoV-2 viral load in the upper respiratory tract of children and adults with early acute COVID-19. Clinical Infectious Diseases 2020;ciaa1157.

29. Rafiei Y, Mello MM. The Missing Piece — SARS-CoV-2 Testing and School Reopening. N Engl J Med 2020;NEJMp2028209.

30. D’Acremont V, Lengeler C, Mshinda H, Mtasiwa D, Tanner M, Genton B. Time To Move from Presumptive Malaria Treatment to Laboratory-Confirmed Diagnosis and Treatment in African Children with Fever. PLoS Med 2009;6(1):e252.

