## Supplementary file for "Antigen rapid tests, nasopharyngeal PCR and saliva PCR to detect SARS-CoV-2: a prospective comparative clinical trial"

### Title: Antigen rapid tests, nasopharyngeal PCR and saliva PCR for the detection of SARS-CoV-2: a prospective comparative clinical trial

*Subjects inclusion and exclusion criteria*

Patients above 18 years were recruited from three different outpatient sites (Unisanté Bugnon, Unisanté Flon, Vidy-Med) and enrolled if they reported having i) at least one major symptom, namely cough, fever, sore throat, anosmia, or ageusia, or ii) a recent close contact with a documented COVID-19 case and presenting with at least one minor symptom (rhinitis, myalgia, headache, fatigue, nausea, vomiting, diarrhoea, abdominal pain, urticaria, vesicules). These criteria corresponded to the regional recommendations for testing (https://coronacheck.ch). Exclusion criteria were unwilling or unable to provide informed consent, or already diagnosed with SARS-CoV-2 in the past, or hospitalized patients, or under anticoagulant therapy.

*Saliva sampling additional details*

Patients collected themselves the saliva for PCR analysis under supervision and according to a video sequence explained step by step by the health professional (see video from 1:14 https://coronavirus.delaware.gov/drive-thru-testing-instructions-for-curative-test-english/). Briefly they swabbed their upper and lower gingival space, plus cheeks on both sides, plus under the tongue to end with the hard and soft palate, and this for at least 20 seconds. They finished by drooling twice liquid saliva in the provided tube, putting the swab in the tube and closing its cap. No coughing or sniffing prior to sample collection was required. Water was avoided 10 minutes prior to collection as much as possible. Other drinks, food, and nasal sprays were avoided 20 minutes before sample collection. The saliva sampling procedure was chosen after multiple attempts in a pilot study in known negative and positive in- and outpatients.

*Detection rates of RDT, NP PCR and saliva PCR*

When considering the 342 (36.9%) patients with a VL $\geq 10^5$ copies/ml by either NP or saliva PCR, the detection rate was 34.2% (31.1-37.3) for RDT, 36.2% (33.1-39.4) for saliva PCR, and 36.6% (33.5-39.8) for NP PCR (p=0.4 and 0.3, respectively).

When considering the 329 patients with a VL $\geq 10^5$ copies/ml by NP PCR, sensitivity of RDT was 94.5% (91.5-96.7%).

*Comparison of virus loads according to RDT result and type, duration of symptom and saliva volume*

Virus loads (by NP PCR) were significantly higher in patients with RDT positive (median $1.9x10^7$; IQR $2.7x10^6$-$1.0x10^8$) than in those with RDT negative ($4.2x10^4$; IQR $2.8x10^3$-$6.2x10^5$) ($p<0.001$). Among patients with an RDT positive, those with a color band of high intensity had also significantly higher virus loads by NP PCR (median $2.8x10^7$; IQR $5.8x10^6$-$1.3x10^8$) than in those with a color band of low intensity (median $7.1x10^5$; IQR $1.5x10^5$-$3.2x10^6$) ($p<0.001$) (Supplementary Figure 3).

Virus loads (by NP PCR) were significantly higher (median $1.3x10^7$; IQR $1.3x10^6$-$8.4x10^7$) in patients with major symptoms than in those with minor symptoms only (median $2.9x10^6$; IQR $8.0x10^2$-$6.6x10^7$) ($p=0.007$), but the number of positive patients with minor symptoms was very small (8). Virus loads (by saliva PCR) was not significantly higher in patients with high volume of saliva (median $1.3x10^5$; IQR $2.2x10^4$-$9.7x10^5$) than those with low volume (median $9.4x10^4$; IQR $9.6x10^3$-$5.1x10^5$) ($p=0.3$) (Supplementary Figure 4). The two PCR were equivalent also in patients with ageusia or sore throat and no other symptom outside anosmia ($p=0.7$).

*Health professional and patient perception*

There was first reluctance of health professionals to change from PCR to RDTs, since they were worried to slow down the flux of patients because of the 15 minutes to wait before reading the test. This issue was addressed with a reorganization, and, actually, the level of satisfaction of the health professionals increased during the study period since they could inform the patient of his/her SARS-CoV-2 status within 15 minutes. The patients were also eager to know the result on site, or just after leaving the center through an SMS.
